# Acronym Disambiguation in Clinical Notes from Electronic Health Records

**DOI:** 10.1101/2020.11.25.20221648

**Authors:** Nicholas B. Link, Selena Huang, Tianrun Cai, Zeling He, Jiehuan Sun, Kumar Dahal, Lauren Costa, Kelly Cho, Katherine Liao, Tianxi Cai, Chuan Hong, in collaboration with the Million Veteran Program

## Abstract

**Objective:** The use of electronic health records (EHR) systems has grown over the past decade, and with it, the need to extract information from unstructured clinical narratives. Clinical notes, however, frequently contain acronyms with several potential senses (meanings) and traditional natural language processing (NLP) techniques cannot differentiate between these senses. In this study we introduce an unsupervised method for acronym disambiguation, the task of classifying the correct sense of acronyms in the clinical EHR notes.

**Methods:** We developed an unsupervised ensemble machine learning (CASEml) algorithm to automatically classify acronyms by leveraging semantic embeddings, visit-level text and billing information. The algorithm was validated using note data from the Veterans Affairs hospital system to classify the meaning of three acronyms: RA, MS, and MI. We compared the performance of CASEml against another standard unsupervised method and a baseline metric selecting the most frequent acronym sense. We additionally evaluated the effects of RA disambiguation on NLP-driven phenotyping of rheumatoid arthritis.

**Results:** CASEml achieved accuracies of 0.947, 0.911, and 0.706 for RA, MS, and MI, respectively, higher than a standard baseline metric and (on average) higher than a state-of-the-art unsupervised method. As well, we demonstrated that applying CASEml to medical notes improves the AUC of a phenotype algorithm for rheumatoid arthritis.

**Conclusion:** CASEml is a novel method that accurately disambiguates acronyms in clinical notes and has advantages over commonly used supervised and unsupervised machine learning approaches. In addition, CASEml improves the performance of NLP tasks that rely on ambiguous acronyms, such as phenotyping.

## INTRODUCTION

A large amount of important clinical data is embedded in the narrative notes within electronic health record (EHR) systems. Mining these EHR data requires processing these unstructured clinical narratives. Current approaches to process unstructured text are mainly based on natural language processing (NLP) techniques [1-6], which have been deployed for tasks such as information extraction, phenotyping [7-14], and making notes more readable [15]. A challenge of applying NLP methods to EHR text is the use of ambiguous acronyms to describe important medical terms. For example, RA is often used as an abbreviation for “rheumatoid arthritis” but it can also represent “room air” or “right atrium”. The use of ambiguous acronyms is ubiquitous – Moon et. al 2012 found 440 acronyms and abbreviations with 949 total senses in a sample of clinical notes [16]. Directly using NLP results without correctly determining what the acronym stands for in context, a process known as acronym disambiguation, hampers the performance of research and clinical applications. As a motivating example, we focus on identifying when the acronyms RA, MS, and MI mean their target senses – “rheumatoid arthritis”, “multiple sclerosis”, and “myocardial infarction” – as these acronyms are highly relevant for information extraction relating to these diseases. For clarity throughout the paper, we will only use “RA”, “MS”, and “MI” to represent the acronyms, not their long forms (i.e. when discussing the diseases/phenotypes we will always use “rheumatoid arthritis” as opposed to “RA”).

Prior methods for acronym disambiguation in biomedical text such as PubMed and WebMD have been developed [17-19]. However, compared to biomedical text, clinical text in EHR provides extra challenges to disambiguation because of its informal and unstructured nature [20] and the more ambiguous use of words [21]. Recent efforts have been devoted to developing supervised acronym disambiguation algorithms for clinical text in EHRs [22]. These approaches rely on chart review labels of the acronyms of interest; although they are generally successful in disambiguation tasks, manually annotating acronyms is time-consuming and is not feasible for large sets of acronyms. As well, the training sample size is limited by the number of annotated labels; it may not be clear ahead of time how many labels are needed to effectively train the algorithm, especially because this number depends on the sense distribution of acronyms [20].

Unsupervised and knowledge-based methods (methods that rely on knowledge sources such as medical dictionaries) alleviate the need for labels. Some unsupervised methods have exploited acronym sense expansions, defined as the fully written out meaning of the acronym in that instance (e.g. “rheumatoid arthritis” or “right atrium” for RA) for semi-supervised or unsupervised disambiguation [23-28]. A limitation of these methods is that they rely on knowing a sense inventory, i.e. a dictionary of possible acronym meanings; however, common inventories can have limited sense coverage. For example, Moon et. al 2014 found that short form – sense pairs matched only 2.3% of the time across three sense inventories [29].

Embedding approaches such as *word2vec* [30] and *GloVE* [31] that use distributed representations of words have improved the accuracy of supervised and unsupervised disambiguation tasks. Finley et. al proposed a semi-supervised method using word embeddings and sense expansion to disambiguate clinical acronyms [25], and Joopudi et. al adapted this method to use neural network-based word embeddings [27]. However, these approaches are still knowledge-based, so they rely on a sense dictionary.

In this paper we propose CASEml (**C**lassifying **A**cronym **S**ense using **E**nsemble **m**achine **l**earning), an unsupervised method for acronym disambiguation in clinical text. Specifically, CASEml is an ensemble (combination) of two models – one using context word embeddings and another leveraging visit-level text and billing information – to identify when an acronym means its target sense. CASEml only relies on the knowledge of one target sense rather than all possible senses. We validated CASEml using note data from the Veteran Affairs (VA) to classify three acronyms: RA, MS, and MI. The results indicate that CASEml can accurately predict acronyms as well or better than state-of-the-art supervised methods in both sets of data. To further analyze the usefulness of CASEml, we evaluated the impact of applying CASEml to medical notes on a downstream NLP task: developing a phenotype algorithm for rheumatoid arthritis using EHR data. We showed that for phenotyping, CASEml outperforms knowledge-based methods and significantly outperforms naïve methods of handling the acronym RA.

## METHODS

### Materials/Data

This study used clinical text and billing codes from the EHR of the Veterans Affairs Healthcare Centers Data for RA and MS were extracted from the Million Veterans Project [32] while data for MI were extracted from the general VA EHR.

For the validation of CASEml there are two types of labels: *acronym-level labels* of RA, MS and MI, and *patient-level labels* of rheumatoid arthritis (i.e., whether the patient has the disease) to evaluate the phenotyping performance. The rheumatoid arthritis phenotype prediction algorithm (discussed below) is only applied to patients with at least one rheumatoid arthritis ICD code, the so-called “filter-positive” set. Since filter-positive patients are often the group of interest for applications of NLP, we performed CASEml separately on notes from the filter-positive set, defined as patients with at least one related ICD code to the target sense/phenotype (multiple sclerosis for MS and myocardial infarction for MI), as well as notes from all patients. For RA and MS, the acronym-level labels were created by randomly sampling 100 notes from filter-positive patients and 100 notes from filter-negative patients; for MI, 100 notes from filter-positive patients who live in Michigan (because here MI often means “Michigan” but outside of the state it almost always means “myocardial infarction”) and 100 notes from filter-positive patients who live in Minnesota as a control group were randomly sampled. For the sampled notes, each acronym was assigned a binary label indicating if the acronym means the target sense. When applying disambiguation methods, we are often only interested in the occurrence of one target sense, so in this study we only aim to classify that one sense. In other words, we treat the disambiguation problem as a binary classification of a target sense rather than a multiclass classification of multiple senses.

The patient-level labels were created by randomly sampling 227 filter-positive patients from MVP and then domain experts conducted chart review and assigned the phenotype labels for rheumatoid arthritis.

### CASEml approach

CASEml is an ensemble (combination) of two models that incorporate different levels of information. The first model, *RFCUI*_*ICD*_, is a visit-level prediction model using random forest (RF) to classify a noisy label representing the target sense; in this case, we used ICD codes related to the target sense as the training label. The second model, *wordvec*_*score*_, uses vector representations of words (word embeddings) to compare the context of an acronym in clinical text to the average context of its target sense. Finally, the predicted probabilities from these two models are averaged to generate final probabilities, which can be converted to binary classifiers using the estimated target sense prevalence. A flow-chart summary of CASEml is shown in Figure 1.

**Figure 1:**
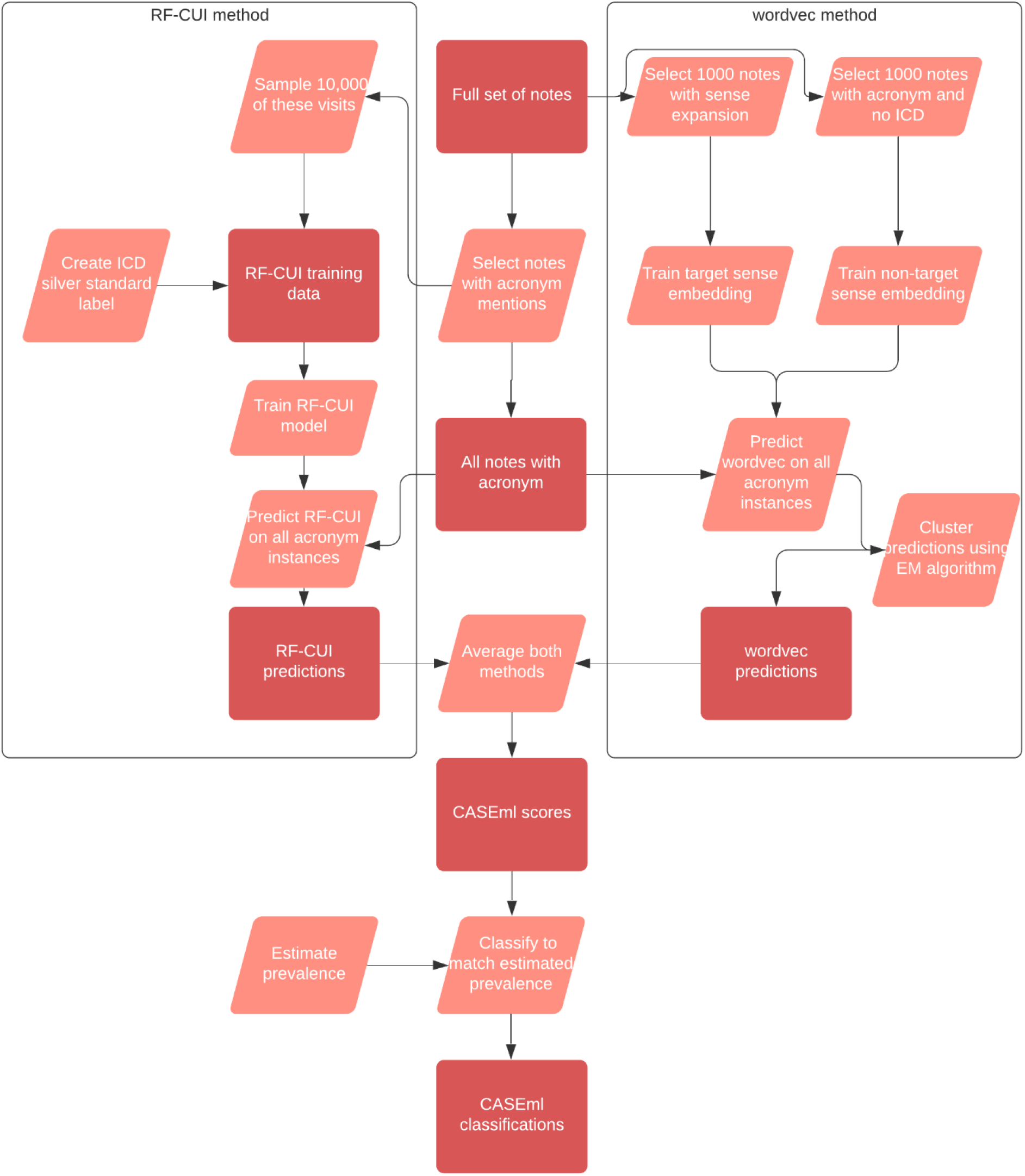
Flowchart of the CASEml method. Rectangular boxes represent data sets and rhombuses represent actions.

### Model 1: *RF* − *CUI*_*ICD*_

The input features for *RF*−*CUI*_*ICD*_ were counts of concept unique identifiers (CUIs), which are groups of terms that have the same meaning, in clinical notes. To generate the feature list, CUIs related to the acronym target sense were extracted from the Unified Medical Language System (UMLS) dictionary [33]. Mentions of these CUIs were extracted from notes with the acronym of interest using NILE, an NLP tool for efficient named entity recognition in EHR text [1]. As well, a set of ICD codes related to the target sense was created [34]. Since the proposed method is unsupervised without using any gold-standard acronym labels, the ICD codes were used to create a binary “silver-standard” (or “machine created”) label for each note. It is possible to use note mentions of the target sense or CUIs related to the target sense to create silver-standard labels, but we chose to use ICD codes in this study.

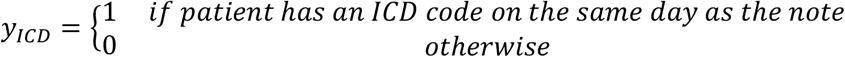

A random forest model to classify *y*_*ICD*_ with the CUI features, called *RF*−*CUI*_*ICD*_, was trained with the randomForest_4.6-14 package in R with default parameters (500 trees, 9 features available at each node for rheumatoid arthritis and 3 for multiple sclerosis) [35].

### Model 2: *wordvec*_*score*_

While *RF*−*CUI*_*ICD*_ predicts the meaning of an acronym using broader visit-level information, often the textual context of an acronym is the most useful predictor of its meaning. To use the acronyms’ contexts, we implemented an unsupervised context embeddings method, *wordvec*_*score*_, to classify a target sense, adapted from methods introduced in [25] and [27].

### Training the word embeddings

The embeddings were taken from previous work [36]; first, the word-context pair cooccurrence was calculated from 1.7 million full text journal articles obtained from PubMed central. Word-context pairs were counted if they appeared within a window of 10 terms from each other, as previous literature has identified that expanding beyond this window size does not generally improve disambiguation performance [37]. A word-context pointwise mutual information (PMI) matrix was then created and factorized to create dimension 500 embeddings. Levy and Goldberg demonstrated that this matrix factorization performs skip-gram with negative sampling as originally introduced by Mikolov et. al [30 38], showing that the vectors obtained are near state-of-the-art.

### Constructing word embeddings with à la carte

À la carte [39] is a method to denoise embeddings *u*_*w*_ by regressing them onto their context embeddings 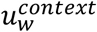, thus making the embeddings more representative of their context in the corpus. The context embeddings were created by equation (1), where *C*_*w*_ is a random set of 1000 contexts of the word *w*. The linear transformation matrix *A* was found by solving equation (2) and then used to transform the embeddings to their denoised form *ν*_*w*_ via equation (3), where *V* is the intersection of words in the set of PMC-based embeddings and in the clinical notes. The words in equation (2) were weighted by the log count of their frequency, as this has been recommended to improve à la carte [39].

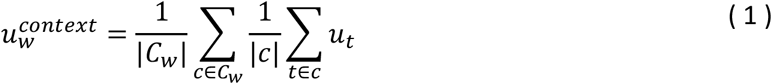

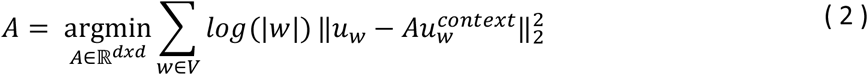

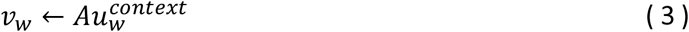

### Training the acronym-context vectors

For each acronym, two embeddings were trained: *ν*_*S*+_, representing the target sense, and *ν*_*S*−_, representing the non-target sense, grouping all other senses together (e.g. “room air” and “right atrium” together for RA). To create *ν*_*S*+_, 1000 instances of the sense expansion in the text were randomly sampled and the weighted average of the context embeddings was taken.

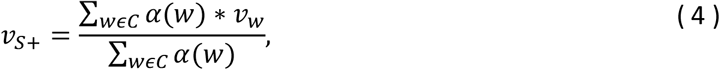

The weights *α*(*w*) amplify words by their inverse frequency via the equation (2), where *tf*_*w*_ is the frequency of term *w* and *α* is the mean word frequency, roughly 4 ∗ 10^−5^.

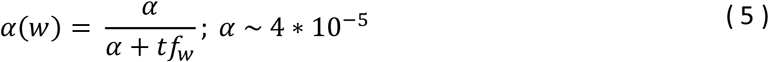

To create *ν*_*S*−_, we randomly sampled 1000 instances of the acronym where there was no related ICD code on the same day, indicating that the acronym likely did not mean the target sense. We then computed the weighted average of the contexts of these instances as was done in equation (4).

### Predicting target senses with embeddings

To fully use the information from both positive and negative samples, we constructed the prediction score by subtracting *cosine*(*ν*_*context*_, *ν*_*S*−_) from *cosine(ν*_*context*_, *ν*_*S*+_). Our preliminary analysis showed that this step significantly outperformed using just the positive sample, *cosine*(*ν*_*context*_, *ν*_*S*+_). As well, we found that performing a logit transformation, adapting for scores from −1 to 1, on the cosine similarities improved the performance.

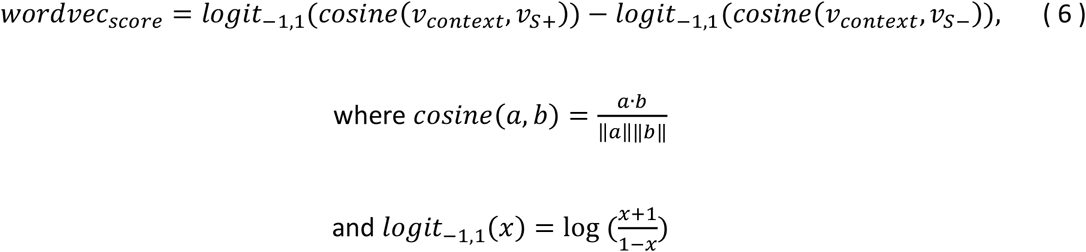

The output of this model is scores of the likelihood of acronyms meaning the target sense; therefore, it is not a direct classification or probability. To convert this score to a probability, we fit a univariate finite Gaussian mixture model using the flexmix package in R [40].

### Ensemble model: CASEml

We averaged the predicted probabilities from *RF*−*CUI*_*ICD*_ and *wordvec*_*score*_ to create a final probability for CASEml. To classify acronym instances, we chose a probability cutoff that resulted in a sense prevalence equal to an estimate of the true sense prevalence. For example, if the estimated true sense prevalence is *prev*, then we determine a cutoff *c* such that *mean*(*CASEml* > *c*) = *prev*. The estimate of prevalence used in this study was the proportion of acronym instances with a CUI disease mention or a disease ICD code on that day.

### Evaluating CASEml

#### Methods for Comparison

Two baseline models were considered for comparison: a standard baseline model, majority frequent sense (MFS), and a knowledge-based method (KB) from Finley et. al [25]. MFS always selects the most frequently occurring sense of an acronym, so in practice it requires manually reviewed labels to know this sense. KB extracts multiple possible senses for an acronym and, similar to CASEml, derives an embedding for each sense from the notes. For each acronym instance, KB computes the cosine similarities of the acronym context embedding with the sense embeddings and classifies the acronym as the sense with the highest cosine similarity. The context is taken with a window of 12 and a sigmoid-weighting function is used to weight closer words more heavily. As well, the terms are weighted by the inverse document frequency and the square root of the embeddings is taken. For RA and MS, as was done in Finley et. al, the possible acronym senses were taken from a previously published knowledge-base [29]; for RA the senses are “rheumatoid arthritis”, “right atrium” and “room air” (which matches the common senses found in this study) and for MS the senses are “multiple sclerosis” and “morphine sulfate” (which misses two of the three most common senses found in this study – “Ms.” And “milliseconds”). MI was not present in the knowledge database, so we used the common acronym senses found in our chart review – “myocardial infarction” and “Michigan”.

#### Acronym-level evaluation

CASEml yields both probabilities and classifications for predicting the target sense of an acronym. We calculated the area under the receiver operating curve (AUC) for the CASEml probabilities and the accuracy for the CASEml, MFS, and KB classifications. Since 100 filter-positive notes and 100 filter-negative notes were used for validation, the results for the full data set were weighted to match the true proportion of filter-positive and filter-negative notes with acronym mentions. If *P* was the percentage of notes with acronym mentions that are filter-positive, then the filter-positive labels were weighted by *P* and the filter-negative labels were weighted by *1-P*. Since equal numbers of notes were labeled from the filter-positive and filter-negative sets, weighting the labels by *P* and *1-P* scaled the results to represent the true proportion of filter-positive and filter-negative notes with acronym mentions. The same scaling was applied to the Michigan + Minnesota data set for MI, where *P* represented the percentage of notes with MI mentions in Michigan out of both states.

#### Acronym WSD for downstream phenotyping prediction

We evaluated the impact of using CASEml for downstream phenotyping prediction of rheumatoid arthritis. Previous studies have found that the patient-level counts of the rheumatoid arthritis CUI, C0003873, are useful for phenotyping rheumatoid arthritis [41]. C0003873 contains terms such as “arthritis, rheumatoid” and “proliferative arthritis” as well as the commonly used, yet ambiguous, acronym “RA”. To perform phenotyping prediction, the acronym classifications obtained by disambiguation approaches were further aggregated to patient-level. We tested four methods to classify instances of RA and included them in the NLP counts of C0003873: (1) always classify RA as negative (not meaning “rheumatoid arthritis”), (2) always classify RA as positive (i.e. use MFS), (3) use KB classifications and (4) use CASEml classifications. More details of the phenotyping process are provided in supplementary section 2.

## RESULTS

### Acronym-level classification

Table 1 shows the sense distributions of the acronyms from the validated notes. In the full data set, the distributions were weighted by the filter-positive acronym percentage *P* used to weight the validation results. *P* is 0.15 for RA, 0.08 for MS, and 0.83 for MI.

**Table 1:** Distribution of acronyms RA, MS, and MI

| Acronym | Full data set |  |  | Filter-positive set |  |  |
| --- | --- | --- | --- | --- | --- | --- |
|  | *Number of notes reviewed* | *Number of acronyms* | *Percentage of senses (weighted to match filter/non-filter distribution)* | *Number of notes reviewed* | *Number of acronyms* | *Percentage of senses* |
| RA | 200 | 311 | Rheumatoid arthritis: 17%<br>Room air: 72%<br>Right Atrium: 11%<br>Other: 0% | 100 | 188 | Rheumatoid arthritis: 69%<br>Room air: 22%<br>Right Atrium: 7 %<br>Other: 2% |
| MS | 200 | 385 | Multiple sclerosis: 12%<br>Ms. (Title): 34%<br>Milliseconds: 30%<br>Other: 24%* | 100 | 215 | Multiple sclerosis: 83%<br>Ms. (Title): 10%<br>Milliseconds: 3 %<br>Other 3%* |
|  | Michigan + Minnesota |  |  | Michigan |  |  |
| MI | 200 | 289 | Myocardial Infarction: 43%<br>Michigan: 54%<br>Other: 3% | 100 | 147 | Myocardial Infarction: 33%<br>Michigan: 65%<br>Other: 3% |

\* "Other" for MS includes "musculoskeletal", "masters of science", and "mental status"

Table 2 compares the accuracy of CASEml, KB, and MFS. The estimated target sense prevalences used to determine the classification cutoffs for CASEml in the full and filter-positive data sets were 13% and 67% for RA, 8% and 73% for MS, and 20% and 22% for MI.

**Table 2:** Accuracy of the three methods on the full data set and the filter-positive set

| Data set | Measure | Acronym | CASEml | KB | MFS* |
| --- | --- | --- | --- | --- | --- |
| Full Data Set | Accuracy | RA | 0.947 | 0.955 | 0.832 |
|  |  | MS | 0.911 | 0.32 | 0.879 |
|  |  | MI | 0.706 | 0.895 | 0.572 |
|  |  | Average | 0.855 | 0.723 | 0.761 |
| Filter Positive | Accuracy | RA | 0.824 | 0.846 | 0.686 |
|  |  | MS | 0.707 | 0.763 | 0.833 |
|  |  | MI | 0.802 | 0.891 | 0.673 |
|  |  | Average | 0.778 | 0.833 | 0.731 |
| *\* The most common senses for the full note set were different from the filter-positive set for both acronyms tested, so MFS had a different classification between both sets.* |  |  |  |  |  |
| *The filter positive set is the Michigan only set for MI and the full data set is the Michigan + Minnesota set. Since MI is not in the knowledge dictionary, KB for MI is a supervised method, relying on manually selecting the possible senses of the acronym. The estimates of accuracy for CASEml are from estimated sense prevalences; using the estimates of the true sense prevalence generally yields more accurate results.* |  |  |  |  |  |

In the full note set, CASEml significantly outperformed MFS, while in the filter-positive set it was more accurate for RA and MI and less accurate for MS. CASEml performed significantly better than KB for MS in the full data set and slightly worse for the remaining sets.

### Rheumatoid arthritis phenotype prediction

Figure 1 shows the receiver operating curves and area under these curves (AUC) for the four methods of rheumatoid arthritis prediction tested. Using CASEml to classify RA returned a phenotyping AUC of 0.937, which is higher than the 0.928 AUC from using KB, and significantly higher than the 0.871 AUC from MFS and 0.91 AUC from excluding RA counts.

**Figure 1:**
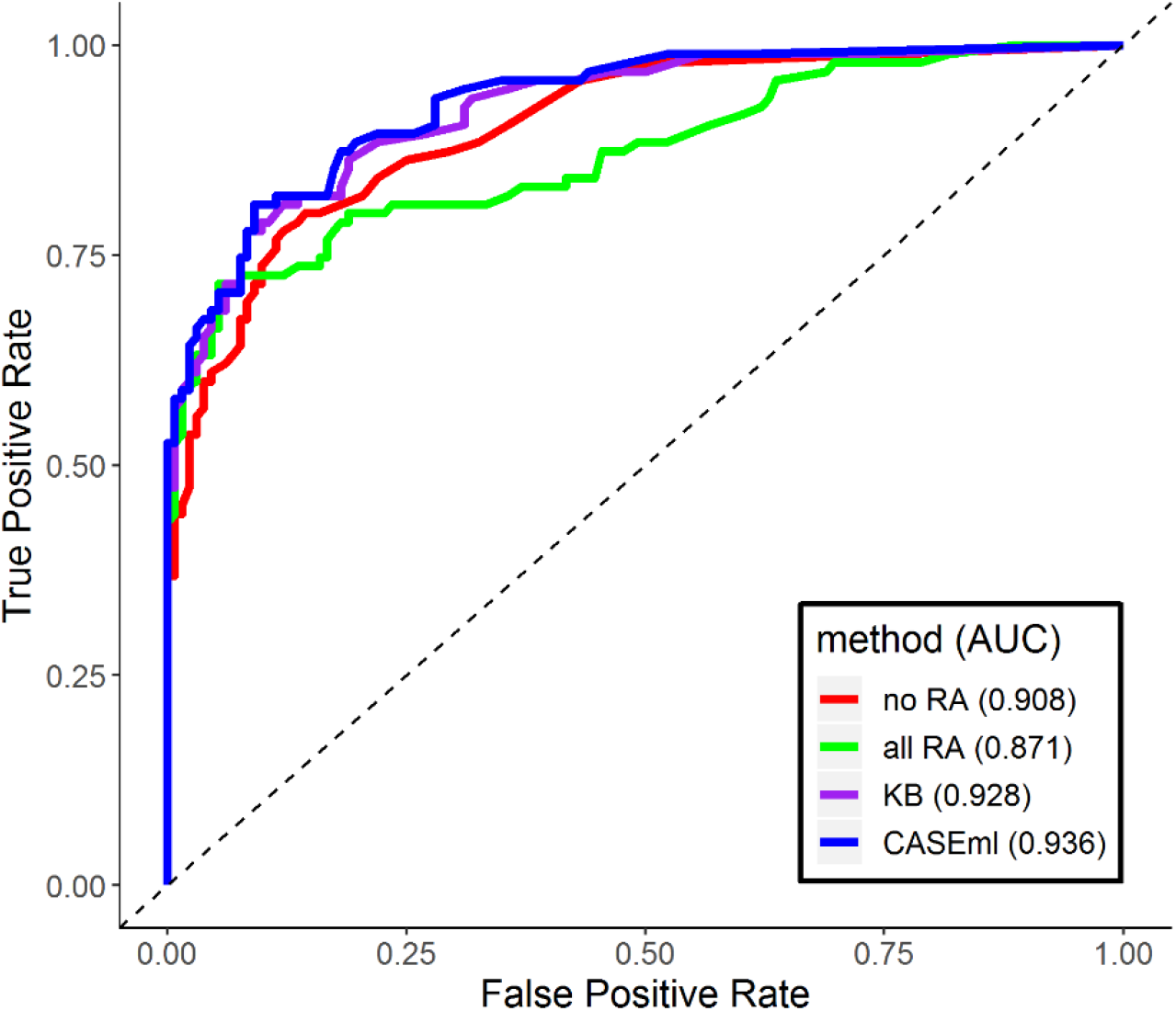
Receiver operating curves (ROC) for rheumatoid arthritis phenotype prediction using NLP. Red: ROC if RA is never included in the NLP feature. Green: RA always counted in the NLP feature. Purple: KB is used to identify when to include RA in the feature. Blue: CASEml is used to identify when to include RA in the feature.

## DISCUSSION

In this study, we developed an unsupervised acronym disambiguation method, CASEml, and demonstrated its effectiveness in disambiguating RA, MS, and MI in clinical EHR notes from 112 VA centers across the United States. CASEml ensembles two approaches: a random forest model using CUI counts trained on silver-standard labels and a model using word embeddings to compare the similarity of the acronym context with the target sense. We created a CASEml classifier by leveraging an estimated acronym sense prevalence. We compared the predictions of CASEml with MFS, a baseline method, and KB, another unsupervised method, and showed that CASEml generally outperforms both. Finally, we demonstrated that CASEml improves the performance of NLP-based phenotyping.

### Analysis of the ensemble method

In many machine learning applications, ensemble models of multiple individually trained models outperform the individual models themselves [42 43]. In this study, *RF*−*CUI*_*ICD*_ and *wordvec*_*score*_ are two base models for CASEml which have different architectures and use different levels of information. Because the models make partially independent and supplementary decisions, as long as neither model is significantly worse than the other, combining them will usually reduce the overall bias in the final result. While *wordvec*_*score*_ outperformed *RF*−*CUI*_*ICD*_ for all three acronyms, on average both models independently performed worse than the ensemble model (Supplementary Table 2).The results indicate that KB is often sufficiently accurate but can be completely inaccurate. For RA and MI, KB was more accurate than CASEml, while for MS, CASEml was significantly more accurate. One reason KB performed poorly for MS because the senses in the knowledge-dictionary are not representative of the true common senses. However, even after manually selecting the most common senses in this set of notes – multiple sclerosis, miss, milliseconds, and musculoskeletal – KB had an accuracy of 0.493 in the filter-positive set and 0.494 in the full data set. This demonstrates the additional benefit of CASEml over KB (and other published unsupervised methods) in that it does not rely on the sense expansion of the non-target senses and it also uses *RF*−*CUI*_*ICD*_, which significantly improved the accuracy and AUC for MS.

If the true target sense prevalence of an acronym is available in practice, it could be used to determine the classification cutoffs, but for the purpose of keeping CASEml unsupervised in this study we used the estimated target sense prevalence from ICD and NLP features. When using the true prevalence of acronym senses for classification, the accuracy of RA, MS, and MI change to 0.983, 0.896, 0.86 for the full data set and 0.824, 0.805, 0.871 for the filter-positive set, an average improvement of .06 for both data sets.

In cases where the context distribution of an acronym sense expansion is different than that of the acronym itself, e.g. if “rheumatoid arthritis” was used differently in text than “RA” representing “rheumatoid arthritis”, *wordvec*_*score*_ might perform poorly (KB would similarly perform poorly). As well, if a visit’s ICD codes are a poor surrogate for the true acronym meaning or if the visit-level CUIs are not predictive of acronym sense, *RF*−*CUI*_*ICD*_ would perform poorly.

### The visit-level model

*RF*−*CUI*_*ICD*_ captures broad information – mentions of related medical concepts – about the visit where an acronym is mentioned. Other studies have used visit-level information, though not extensively [15]. To the best of our knowledge, this is the first study to use visit-level concept counts as features or structured billing codes to create silver-standard labels. However, *RF*−*CUI*_*ICD*_ was generally less accurate than *wordvec*_*score*_, signifying that the immediate context of an acronym is more indicative of its sense than the broader information and that *RF*−*CUI*_*ICD*_ would not be good as a stand-alone method.

We compared random forest, logistic and lasso regression models in this approach, and the random forest model performed slightly better on the validation set, though there was not a significant difference in the results.

### Word embeddings and à la carte

As reported in previous work [25 27], the unsupervised (or semi-supervised) word embeddings approach using sense expansions accurately disambiguated acronyms. An additional step used in this study was à la carte, which reduces noise and widens the difference between unrelated word embeddings; in fact, the average cosine similarity of two random embeddings in *V* decreased from 0.887 before applying à la carte to 0.449 after. Previous work has found that acronym sense distributional similarity (how closely related acronym sense embeddings are) hurts the performance of disambiguation tasks [20]; à la carte may alleviate this issue.

### Acronym disambiguation and downstream NLP tasks

We demonstrated that acronym disambiguation with CASEml can significantly improve the performance of NLP-based phenotyping. Phenotyping in EHR is important for cohort creation, creating co-morbidities for other studies, and running genetic analyses. To the best of our knowledge, this is the first study to show that acronym disambiguation can improve the performance of phenotyping tasks. While we only explored one application to phenotyping, there are many other acronyms that could be important to analyze for phenotyping tasks, e.g. ED for eating disorders and erectile disfunction.

### Limitations

While CASEml does not require manually annotated labels, it does require some manual input. For each acronym, the target sense needs to be identified, and for this target sense a list of ICD codes and related CUIs needs to be created. In this paper we only considered when the target sense of an acronym is a disease, which makes the list of related ICD codes and CUIs easy to obtain. In other cases, when the target sense is not a disease or well-defined clinical concept (such as “milliseconds” for MS), using CASEml, specifically *RF*−*CUI*_*ICD*_, could be problematic.

Another limitation of CASEml is that it is designed specifically for binary classification of a target sense. It is not directly applicable to disambiguation of multiple acronym senses, though the methods presented in this study could be adapted to that type of problem.

### Future work

To analyze the robustness of CASEml, future directions include applying this approach to a broader set of medical acronyms. It is possible the finite mixture model would not converge for acronyms with a highly dominant sense. As well, since only the phenotype label for rheumatoid arthritis was available at the time of this study, CASEml and other disambiguation methods should be tested on acronyms affecting other phenotypes; this would allow for a better comparison of CASEml, KB, and MFS.

For *RF*−*CUI*_*ICD*_, silver-standard labels other than *y*_*ICD*_ could be tested, and it would be useful if these silver-standard labels did not require any manual curation to create. Another improvement of CASEml could be for *wordvec*_*score*_ to use word embeddings created from the clinical text as opposed to biomedical text.

## CONCLUSION

In this study we presented the unsupervised Classification of Acronym Sense with Ensemble machine learning (CASEml) method to classify when an acronym means a target sense. We demonstrated the usefulness of combining two approaches: a visit-level random forest model using billing codes and CUI counts and a context-driven word embeddings model with a denoising step, à la carte. Further, we demonstrated that CASEml improves downstream NLP-based phenotype prediction. CASEml is able to efficiently and accurately disambiguate acronyms, thus improving our ability to extract useful information from clinical text in the EHR.

## Data Availability

Due to the sensitive nature of the data, it is not possible to share.

