## Supplementary Materials for "Acronym Disambiguation in Clinical Notes from Electronic Health Records"

### Supplementary Section 1: Confusion matrices for $y_{ICD}$

The confusion matrices for CASEml, KB, and the silver-standard label $y_{ICD}$ with the true labels are given in supplementary tables 1 and 2. $y_{ICD}$ is highly specific (0.998, 0.995, and 0.974 for RA, MS, and MI in the full data set and 0.984, 0.778, and 0.96 in the filter-positive set) but not sensitive (0.398, 0.413, 0.263 in the full data set and 0.472, 0.592, 0.271 in the filter-positive set).

**Supplementary Table 1: Confusion Matrices (proportion rather than raw counts) – Full data Set**

| **Classification** | **RA** | | | **MS** | | |
|---|---|---|---|---|---|---|
| $y_{ICD}$ | | Label = 1 | Label = 0 | | Label = 1 | Label = 0 |
| | Class = 1 | 0.067 | 0.001 | Class = 1 | 0.05 | 0.004 |
| | Class = 0 | 0.101 | 0.831 | Class = 0 | 0.071 | 0.875 |
| CASEml | | Label = 1 | Label = 0 | | Label = 1 | Label = 0 |
| | Class = 1 | 0.115 | 0 | Class = 1 | 0.05 | 0.018 |
| | Class = 0 | 0.053 | 0.832 | Class = 0 | 0.071 | 0.861 |
| KB | | Label = 1 | Label = 0 | | Label = 1 | Label = 0 |
| | Class = 1 | 0.144 | 0.021 | Class = 1 | 0.106 | 0.665 |
| | Class = 0 | 0.024 | 0.812 | Class = 0 | 0.015 | 0.214 |

**Supplementary Table 2: Confusion Matrices (proportion rather than raw counts) – Filter-positive data set**

| Classification | RA | | | MS | | |
|---|---|---|---|---|---|---|
| $y_{\mathrm{ICD}}$ | | Label = 1 | Label = 0 | | Label = 1 | Label = 0 |
| | Class = 1 | 0.324 | 0.005 | Class = 1 | 0.493 | 0.037 |
| | Class = 0 | 0.362 | 0.309 | Class = 0 | 0.34 | 0.13 |
| CASEml | | Label = 1 | Label = 0 | | Label = 1 | Label = 0 |
| | Class = 1 | 0.516 | 0.005 | Class = 1 | 0.558 | 0.019 |
| | Class = 0 | 0.17 | 0.309 | Class = 0 | 0.274 | 0.149 |
| KB | | Label = 1 | Label = 0 | | Label = 1 | Label = 0 |
| | Class = 1 | 0.569 | 0.037 | Class = 1 | 0.735 | 0.14 |
| | Class = 0 | 0.117 | 0.277 | Class = 0 | 0.098 | 0.028 |

**Supplementary Section 2: Phenotyping Rheumatoid Arthritis**

Rheumatoid arthritis phenotyping was done in the VA MVP cohort. Only patients with at least one ICD code for rheumatoid arthritis are considered. Out of this group, 227 patients were randomly selected to be chart reviewed, and these are the gold standard labels used in this study. For the sake of comparison, the only feature used in this study to predict the labels is the NLP count of C0003873 without “RA” + classifications of “RA”, though in practice ICD codes are also useful predictors of the rheumatoid arthritis phenotype [41]. The NLP counts are log-transformed, so that the model is

$$label_{RA} \sim \log\left(1 + C0003873_{excluding\ "RA"} + \text{disambiguated "RA"}\right) \quad (1)$$

To get the AUC scores for each disambiguation method, we perform 10-fold cross-validation and average the AUCs across all held-out test sets.

**Supplementary Section 3: AUC of $RF - CUI_{ICD}$, $wordvec_{score}$, and CASEml**

Supplementary table 3 shows that $wordvec_{score}$ always has a higher AUC than $RF - CUI_{ICD}$ – on average 0.077 higher in the full data set and 0.073 in the filter-positive set. However, CASEml usually performs better than $wordvec_{score}$; for RA and MS it had a higher AUC while for MI the AUC was lower. On average, the AUC of CASEml was 0.029 higher than $wordvec_{score}$ in the full data set and 0.058 higher in the filter-positive data set.

**Supplementary Table 3:** AUC of CASEml and the two component models on the full data set and the filter positive set.

| **Data set** | **Measure** | **Acronym** | **CASEml** | $RF - CUI_{ICD}$ | $wordvec_{score}$ |
|---|---|---|---|---|---|
| **Full Data Set** | AUC (95% confidence interval) | *RA* | 0.995 ( , ) | 0.938 | 0.993 |
| | | *MS* | 0.854 | 0.713 | 0.803 |
| | | *MI* | 0.931 | 0.813 | 0.954 |
| | | *Average* | 0.927 | 0.821 | 0.898 |
| **Filter Positive Set** | AUC (95% confidence interval) | *RA* | 0.985 | 0.852 | 0.977 |
| | | *MS* | 0.847 | 0.715 | 0.754 |
| | | *MI* | 0.939 | 0.812 | 0.959 |
| | | *Average* | 0.924 | 0.793 | 0.866 |
